# Dose-dependent oral glucocorticoid cardiovascular risk in people with immune-mediated inflammatory diseases

**DOI:** 10.1101/2020.03.11.20034157

**Authors:** Mar Pujades-Rodriguez, Ann W Morgan, Richard M Cubbon, Jianhua Wu

**Affiliations:** Leeds Institute of Health Sciences, School of Medicine, University of Leeds, Leeds United Kingdom; Leeds Institute of Cardiovascular and Metabolic Medicine, School of Medicine, University of Leeds, Leeds United Kingdom; NIHR Biomedical Research Centre, Leeds Teaching Hospitals NHS Trust, Chapel Allerton Hospital, Leeds, Leeds United Kingdom; School of Dentistry, University of Leeds, Leeds United Kingdom

**Author notes:** **Corresponding author:** Mar Pujades-Rodriguez, Leeds Institute of Health Sciences, University of Leeds, Clarendon Way, Leeds, LS2 9NL, United Kingdom. These authors equally contributed to this work (Mar Pujades-Rodriguez, Jianhua Wu).

**Keywords:** Adverse events, Cohort studies, Dose-response assessment, Giant cell arteritis, Glucocorticoid toxicity, Cardiovascular diseases, Inflammatory bowel disease, Polymyalgia rheumatica, Population, Rheumatoid arthritis, Systemic lupus erythematosus, Vasculitis

## Abstract

**Background:** Evidence for the association between glucocorticoid dose and cardiovascular risk is weak for moderate and low doses. To quantify glucocorticoid dose-dependent cardiovascular risk in people with six immune-mediated inflammatory diseases.

**Methods and Findings:** Population-based cohort analysis of medical records from 389 primary care practices contributing data to the UK Clinical Practice Research Datalink, linked to hospital admissions and deaths in 1998-2017. There were 87,794 patients with giant cell arteritis and/or polymyalgia rheumatica (n=25,581), inflammatory bowel disease (n=27,739), rheumatoid arthritis (n=25,324), systemic lupus erythematosus (n=3951), and/or vasculitis (n=5199); and no prior cardiovascular disease (CVD). Mean age was 56 years and 34.1% were men. Median follow-up time was 5.0 years. Time-variant daily and cumulative glucocorticoid prednisolone-equivalent dose-related risks and hazard ratios of first all-cause and type-specific CVD.

We found 13,426 (15.3%) people with incident CVD, including 6,013 atrial fibrillation, 7,727 heart failure and 2,809 acute myocardial infarction events. At 1 and 5 years, the cumulative risks of all-cause CVD increased from 1.5% in periods of non-use to 9.1% for a daily prednisolone-equivalent dose of ≥25.0mg, and from 7.6% to 29.9%, respectively. We found strong dose-dependent estimates for all immune-mediated diseases (hazard ratio [HR] for <5.0mg daily dose vs. non-use=1.74, 95%CI 1.64-1.84; range 1.52 for polymyalgia rheumatica and/or giant cell arteritis to 2.82 for systemic lupus erythematosus), all cardiovascular outcomes, regardless of disease activity level. The highest estimates were for heart failure and acute myocardial infarction.

**Conclusions:** We estimated glucocorticoid dose-dependent cardiovascular risk in six immune-mediated diseases. Results highlight the importance of prompt and regular monitoring of cardiovascular risk and use of primary prevention treatment at all glucocorticoid doses.

## INTRODUCTION

Patients with immune-mediated inflammatory diseases often receive long-term courses of oral glucocorticoids to reduce disease activity and inflammation during the initial episode and subsequent episodic flares. Prolonged glucocorticoid treatment often causes adverse events, including cardiovascular diseases (CVDs)[1-3]. Glucocorticoids can increase cardiovascular risk through direct and indirect metabolic syndrome enhancement[4-6] and mineralocorticoid effects, including cellular membrane electrolyte-mediated efflux[7-9]. However, the anti-inflammatory and immune-suppressive effects of glucocorticoids could also lower or neutralise the atherosclerotic and vascular injury effects of chronic inflammatory diseases[10]. Demonstration of cost-effectiveness of newly licenced glucocorticoid-sparing drugs, such as biologics, is critical to guide their introduction for the treatment of immune-mediated inflammatory diseases in routine healthcare. These studies require the accurate estimation of glucocorticoid dose-response relationships to quantify cost-savings associated with the toxicity profile of new drugs.

Evidence of the relationship between glucocorticoids and CVDs comes primarily from studies of associations with current baseline medication use or dose[2, 3, 7, 11-16], ignoring the doses previously administered and their changes over time, as well as the concomitant use of other common medications that can affect the risk of CVDs (e.g. non-steroidal anti-inflammatory drugs). Many also have failed to adjust for important cardiovascular risk factors, such as smoking[1, 2, 7, 13, 16]. These studies have reported a dose-dependent risk of CVD with weaker associations for daily prednisolone-equivalent doses lower than 5-10mg[2, 3, 13, 17].

Our study aimed to estimate daily and cumulative dose-dependent oral glucocorticoid cardiovascular disease risk accurately in people diagnosed with six common immune-mediated inflammatory diseases in England.

## METHODS

### Setting and data sources

We analysed linked electronic health records from people registered at family practices in the Clinical Practice Research Datalink (CPRD) between 1^st^ Jan 1998 and 15^th^ March 2017. CPRD contains demographic and lifestyle data, diagnoses (e.g. stroke), prescribed medication and results of laboratory tests and clinical examinations, prospectively recorded during primary care contacts[18].

Previous validation studies have provided evidence of the accuracy of diagnostic and prescribing data[18]. Patients are broadly representative of the UK population with regard to age, sex and ethnicity[18]. CPRD data were linked to hospital records and the mortality registry (Text 1 in S1 File). Hospital records from the Hospital Episode Statistics (HES) (www.hscic.gov.uk/hes) contain diagnoses recorded during elective and emergency hospital admission across all National Health Service hospitals in England. Mortality data from the Office of National Statistics (ONS) (https://www.ons.gov.uk/atoz?query=mortality&size=10) were used to identify dates and causes of death.

### Ethical considerations

The study was approved by the Independent Scientific Advisory Committee for Medicines and Healthcare products Regulatory Agency database research (ISAC), reference 16_146.

### Study design and follow-up

This was a cohort study including all patients continuously registered in a CPRD practice for 1 year or more, aged ≥18 years and free of CVD, who had been diagnosed with at least one of 6 immune-mediated inflammatory diseases commonly treated with oral glucocorticoids at or before the start of follow-up. These were polymyalgia rheumatica, giant cell arteritis (GCA), systemic lupus erythematosus, rheumatoid arthritis, vasculitis, and inflammatory bowel disease (Figure 1 in S1 File). Diagnostic codes used to identify patients with each immune-mediated inflammatory disease are shown in Table 1 in S1 File. For each patient, the follow-up started when they first became eligible. It ended on the earliest of the following dates: occurrence of the outcome analysed (e.g. stroke), leaving the family practice, death, or last data collection date.

**Table 1.** Patient baseline characteristics by type of immune-mediated inflammatory disease.

|  | All immune-mediated diseases | Polymyalgia rheumatica and/or giant cell arteritis | Inflammatory bowel disease | Rheumatoid arthritis | Systemic lupus erythematosus | Vasculitis |
| --- | --- | --- | --- | --- | --- | --- |
|  | N = 87,794 | N = 25,581 | N = 27,739 | N = 25,324 | N = 3,951 | N = 5,199 |
| **Follow-up time (years), mean (SD)** | 5.9 (4.7) | 5.6 (4.3) | 6.4 (5.0) | 6.2 (4.8) | 6.2 (4.9) | 5.5 (4.4) |
| Total | 521,161 | 142,216 | 176,547 | 156,846 | 24,567 | 28,343 |
| Since first recorded disease code | 9.6 (8.7) | 6.8 (5.3) | 11.7 (9.9) | 10.6 (9.4) | 10.3 (8.4) | 7.7 (6.7) |
| **Sociodemographic information** |  |  |  |  |  |  |
| *Males*, n (%) | 29,935 (34.1) | 7,210 (28.2) | 12,992 (46.8) | 6,963 (27.5) | 599 (15.2) | 2,171 (41.8) |
| *Age (years)*, mean (SD) | 56.0 (18.3) | 71.08 (10.9) | 43.9 (16.3) | 56.7 (15.1) | 46.4 (14.9) | 49.6 (18.1) |
| *Ethnicity*, n (%) |  |  |  |  |  |  |
| White | 75,569 (86.1) | 22,371 (87.5) | 24,493 (88.3) | 21,478 (84.8) | 2,942 (74.5) | 4,285 (82.4) |
| Asian | 2,736 (3.1) | 376 (1.5) | 1,025 (3.7) | 855 (3.4) | 278 (7.0) | 202 (3.9) |
| Black | 1,042 (1.2) | 142 (0.6) | 251 (0.9) | 345 (1.4) | 238 (6.0) | 66 (1.3) |
| Other | 1,108 (1.3) | 158 (0.6) | 443 (1.6) | 312 (1.2) | 129 (3.3) | 66 (1.3) |
| *Index of multiple deprivation*, n (%) |  |  |  |  |  |  |
| 1st (least deprived) | 15,943 (18.2) | 5,080 (19.9) | 5170 (18.6) | 4,042 (16.0) | 662 (16.8) | 989 (19.0) |
| 5th (most deprived) | 14,356 (16.4) | 3,180 (12.4) | 4,583 (16.5) | 4,945 (19.5) | 809 (20.5) | 839 (16.1) |
| **Biomarkers**, mean (SD) |  |  |  |  |  |  |
| Body Mass Index (kg/m<sup>2</sup>) | 26.8 (5.8) | 27.5 (5.6) | 25.7 (5.6) | 27.5 (6.1) | 26.2 (5.6) | 27.8 (6.6) |
| C-reactive protein (mg/L) | 24.7 (39.0) | 33.9 (44.5) | 19.2 (38.4) | 19.6 (30.4) | 7.8 (13.6) | 19.6 (40.2) |
| Erythrocyte sedimentation rate (mm/h) | 31.2 (27.0) | 40.2 (29.0) | 20.2 (20.9) | 27.8 (24.2) | 21.7 (22.0) | 21.5 (24.1) |
| Total cholesterol (mmol/L) | 5.2 (1.2) | 5.2 (1.2) | 5.0 (1.1) | 5.2 (1.1) | 5.1 (1.2) | 5.1 (1.2) |
| HDL cholesterol (mmol/L) | 1.5 (0.5) | 1.5 (0.5) | 1.4 (0.4) | 1.4 (0.5) | 1.5 (0.5) | 1.4 (0.5) |
| LDL cholesterol (mmol/L) | 3.06 (1.0) | 3.07 (1.0) | 3.0 (1.0) | 3.10 (1.1) | 3.05 (1.0) | 3.06 (1.1) |
| Haemoglobin (mmol/L) | 13.1 (1.5) | 12.9 (1.4) | 13.3 (1.7) | 13.0 (1.5) | 13.0 (1.5) | 13.5 (1.5) |
| Creatinine (mmol/L) | 81.0 (26.0) | 83.6 (24.5) | 80.0 (21.0) | 77.7 (21.7) | 79.0 (41.0) | 88.5 (55.6) |
| Systolic blood pressure (mmHg) | 134 (19.3) | 141 (18.1) | 126 (17.5) | 134 (18.9) | 127 (18.3) | 132 (18.5) |
| Diastolic blood pressure (mmHg) | 78 (10.1) | 79 (9.9) | 76 (10.1) | 79 (10.2) | 77 (10.3) | 78 (10.4) |
| **Health behaviour** |  |  |  |  |  |  |
| ***Smoking status*, n (%)** |  |  |  |  |  |  |
| Non-smoker | 32,995 (37.6) | 11,559 (45.2) | 9,361 (33.7) | 8,849 (34.9) | 1,201 (30.4) | 2,025 (38.9) |
| Ex-smoker | 11,654 (13.3) | 4,791 (18.7) | 3,079 (11.1) | 2,963 (11.7) | 243 (6.2) | 578 (11.1) |
| Current smoker | 21,264 (24.2) | 5,183 (20.3) | 6,630 (23.9) | 7,222 (28.5) | 1,000 (25.3) | 1,229 (23.6) |
| ***No. of hospitalisation in last year*, mean (SD)** | 0.60 (2.8) | 0.40 (1.90) | 0.77 (2.8) | 0.51 (1.5) | 0.72 (4.2) | 0.92 (7.2) |
| **Comorbidities, n (%)** |  |  |  |  |  |  |
| Diabetes | 5,602 (6.4) | 2,351 (9.2) | 1,107 (4.0) | 1,596 (6.3) | 181 (4.6) | 367 (7.1) |
| Hypertension | 22,072 (25.1) | 10,810 (42.3) | 3,251 (11.7) | 5,960 (23.5) | 736 (18.6) | 1,315 (25.3) |
| Asthma | 12,858 (14.6) | 3,597 (14.1) | 4,322 (15.6) | 3,567 (14.1) | 492 (12.5) | 880 (16.9) |
| Chronic obstructive pulmonary disease | 2,950 (3.4) | 1,280 (5.0) | 498 (1.8) | 999 (3.9) | 59 (1.5) | 114 (2.2) |
| Cancer | 5,005 (5.7) | 2,259 (8.8) | 998 (3.6) | 1,293 (5.1) | 154 (3.9) | 301 (5.8) |
| Renal disease | 1,095 (1.2) | 556 (2.2) | 171 (0.6) | 247 (1.0) | 36 (0.9) | 85 (1.6) |
| **Prescribed medication at baseline, n (%)** |  |  |  |  |  |  |
| Statins | 8,516 (9.7) | 4,437 (17.3) | 1,187 (4.3) | 2,198 (8.7) | 235 (5.9) | 459 (8.8) |
| Any blood pressure lowering medication | 23,755 (27.1) | 11,495 (44.9) | 3,645 (13.1) | 6,445 (25.5) | 802 (20.3) | 1,368 (26.3) |
| Angiotensin converting enzyme inhibitors | 8,151 (9.3) | 4,075 (15.9) | 1,215 (4.4) | 2,145 (8.5) | 226 (5.7) | 490 (9.4) |
| Angiotensin II receptor antagonist | 3,633 (4.1) | 1,960 (7.7) | 471 (1.7) | 853 (3.4) | 118 (3.0) | 231 (4.4) |
| **Prescribed medication in last year, n (%)** |  |  |  |  |  |  |
| Oral glucocorticoids | 14,356 (16.4) | 4,594 (18.0) | 4,583 (16.5) | 4,945 (19.5) | 809 (20.5) | 839 (16.1) |
| Inhaled or nasal glucocorticoids | 13,331 (15.2) | 4,523 (17.7) | 3,581 (12.9) | 3,895 (15.4) | 474 (12.0) | 858 (16.5) |
| Intramuscular or intra-articular glucocorticoids | 1,216 (1.4) | 470 (1.8) | 121 (0.4) | 584 (2.3) | 22 (0.6) | 19 (0.4) |
| Rectal glucocorticoids | 5,800 (6.6) | 531 (2.1) | 4,601 (16.6) | 491 (1.9) | 61 (1.5) | 116 (2.2) |
| Topical glucocorticoids | 2,094 (2.4) | 647 (2.5) | 615 (2.2) | 525 (2.1) | 139 (3.5) | 168 (3.2) |
| Non-steroidal anti-inflammatory drugs | 39,690 (45.2) | 13,998 (54.7) | 5,143 (18.5) | 17,694 (69.9) | 1,371 (34.7) | 1,484 (28.5) |
| DMARDs ever during follow-up | 18,877 (21.5) | 1,112 (4.3) | 4,072 (14.7) | 12,147 (48.0) | 1,234 (31.2) | 316 (6.1) |

Note: DMARDs, disease modifying anti-rheumatic drugs; SD, standard deviation. Data on ethnicity, body mass index, c-reactive protein, erythrocyte sedimentation rate, total cholesterol, LDL-cholesterol, HDL-cholesterol, creatinine, systolic blood pressure, and smoking status were missing for 8.4%, 59.1%, 68.4%, 58.2%, 74.9%, 80.4%, 84.4%, 47.7%, 36.6%, and 24.9% of patients, respectively.

**Fig 1.**
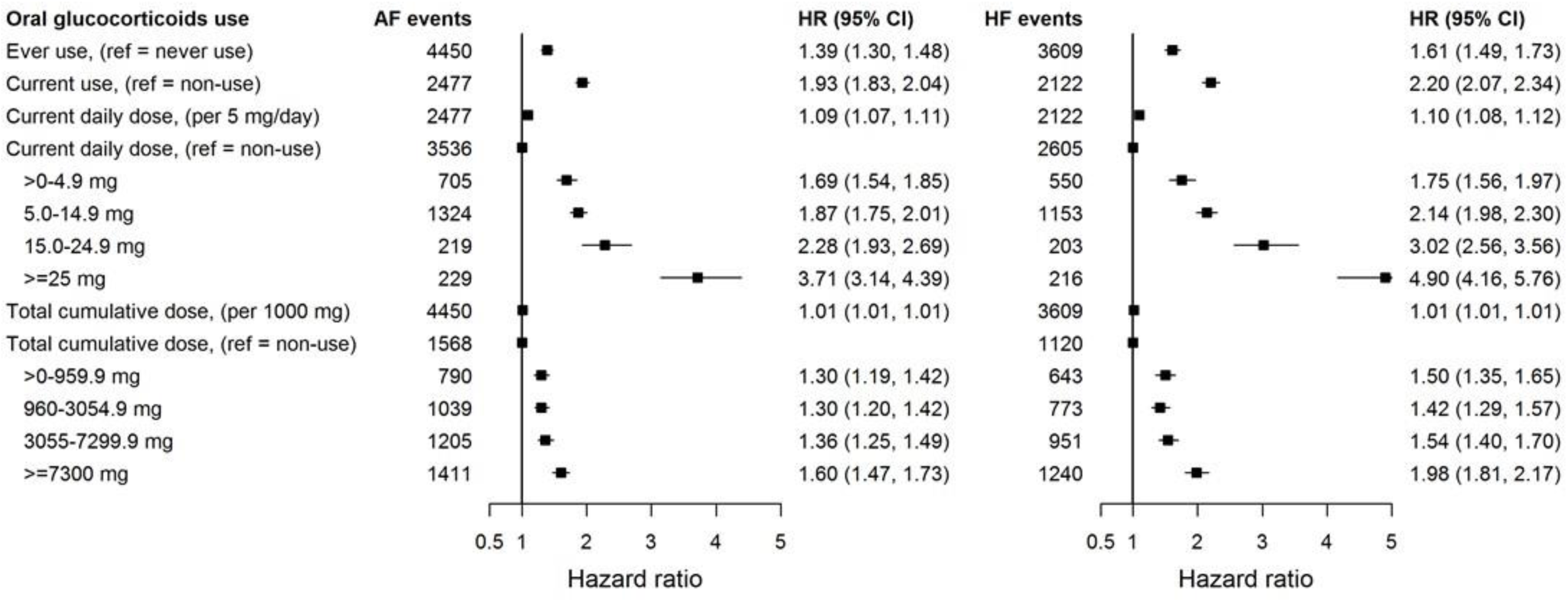
Associations between time variant oral glucocorticoid prednisolone-equivalent dose and incident atrial fibrillation and heart failure for patients with 6 immune-mediated inflammatory diseases. Note: AF, atrial fibrillation; CI, confidence interval; HF, heart failure; HR, Hazard ratios from Cox proportional imputed models adjusted for baseline age, sex, index of multiple deprivation, smoking status, ethnicity, body mass index, type of immune-mediated inflammatory disease, comorbidities (diabetes, diagnosed hypertension, cancer, asthma, chronic obstructive pulmonary disease, and renal disease), biomarkers (total cholesterol, high-density lipoprotein cholesterol, low-density lipoprotein cholesterol, c-reactive protein, creatinine), number of hospital admissions in last year, and prescribed non-oral glucocorticoids; and time-variant use of disease-modifying anti-rheumatic drugs and non-steroidal anti-inflammatory drugs; the practice identifier was included as a random intercept to account for clustering effect.

### Oral glucocorticoid exposure

For each prescription of oral glucocorticoids issued to the patients between one year before the start and the end of the follow-up dates, we derived the daily dose from the recorded product name, which included information on product strength (e.g. 2mg), directions given (e.g. one tablet once a day) and quantity prescribed (e.g. 28 tablets). We then estimated the duration of each oral glucocorticoid prescription dividing the quantity of tablets prescribed by the daily dose. Given the variation in relative anti-inflammatory effects of different types of glucocorticoids, for each prescription we finally converted the daily dosage into milligrams of prednisolone-equivalent dose (Table 2 in S1 File).

**Table 2.** Associations between time variant oral glucocorticoid prednisolone-equivalent dose and incident all-cause cardiovascular diseases by immune-mediated inflammatory disease

|  | Adjusted hazard ratios with 95% CI |  |  |  |  |  |
| --- | --- | --- | --- | --- | --- | --- |
|  | All immune-mediated diseases* | Polymyalgia rheumatica and/or giant cell arteritis | Inflammatory bowel disease | Rheumatoid arthritis | Systemic lupus erythematosus | Vasculitis |
| **No. of events** | 13,426 | 6,267 | 1,937 | 4,236 | 375 | 611 |
| **Ever use** (ref: no used since 1 year prior to follow-up start) | 1.46 (1.40-1.53) | 1.25 (1.15-1.37) | 1.39 (1.26-1.52) | 1.63 (1.52-1.73) | 1.69 (1.35- 2.12) | 1.52 (1.28-1.81) |
| **Current use** (ref: non-use) | 1.95 (1.87-2.02) | 1.69 (1.60-1.78) | 2.71 (2.43-3.02) | 2.11 (1.98-2.25) | 2.56 (2.02- 3.25) | 2.07 (1.71-2.50) |
| **Current daily dose per 5 mg/day** | 1.09 (1.07-1.11) | 1.17 (1.15-1.19) | 1.08 (1.06-1.09) | 1.28 (1.25-1.31) | 1.28 (1.20- 1.37) | 1.19 (1.13-1.25) |
| **Current daily dose** (ref: non-use) | 1.00 | 1.00 | 1.00 | 1.00 | 1.00 | 1.00 |
| 1-4.9 mg | 1.69 (1.57-1.81) | 1.50 (1.37-1.64) | 2.16 (1.66-2.82) | 1.84 (1.62-2.10) | 2.81 (1.92- 4.11) | 1.92 (1.35-2.74) |
| 5.0-14.9 mg | 1.89 (1.81-1.98) | 1.70 (1.59-1.82) | 2.39 (2.03-2.82) | 2.00 (1.85-2.15) | 2.19 (1.62- 2.94) | 1.92 (1.51-2.44) |
| 15.0-24.9 mg | 2.38 (2.13-2.67) | 2.07 (1.80-2.38) | 3.44 (2.49-4.74) | 2.79 (2.21-3.51) | 2.61 (1.15- 5.94) | 2.46 (1.53-3.96) |
| ≥25 mg | 3.64 (3.28-4.04) | 2.76 (2.30-3.32) | 4.35 (3.43-5.52) | 4.98 (4.11-6.03) | 5.66 (3.20-10.01) | 3.07 (1.92-4.91) |
| **Total cumulative dose per 1000 mg** | 1.01 (1.01-1.01) | 1.02 (1.01-1.02) | 1.01 (1.01-1.01) | 1.02 (1.02-1.03) | 1.03 (1.01- 1.04) | 1.03 (1.02-1.04) |
| **Total cumulative dose** (ref: non-use) | 1.00 | 1.00 | 1.00 | 1.00 | 1.00 | 1.00 |
| 1-959.9 mg | 1.37 (1.29-1.45) | 1.24 (1.11-1.38) | 1.21 (1.06-1.38) | 1.47 (1.34-1.61) | 1.65 (1.20- 2.26) | 1.54 (1.23-1.93) |
| 960-3,054.9 mg | 1.35 (1.28-1.43) | 1.14 (1.04-1.26) | 1.35 (1.18-1.54) | 1.52 (1.36-1.68) | 1.40 (0.95- 2.06) | 1.31 (0.95-1.82) |
| 3,055-7,299.9 mg | 1.44 (1.36-1.52) | 1.21 (1.10-1.33) | 1.45 (1.24-1.69) | 1.72 (1.55-1.90) | 1.60 (1.10- 2.34) | 1.27 (0.94-1.70) |
| ≥7,300 mg | 1.76 (1.66-1.86) | 1.50 (1.36-1.66) | 1.85 (1.58-2.16) | 1.80 (1.65-1.97) | 2.09 (1.53- 2.85) | 1.91 (1.49-2.45) |

Note: CI, confidence interval; Hazard ratios from Cox proportional imputed models adjusted for baseline age, sex, index of multiple deprivation, smoking status, ethnicity, body mass index, comorbidities (diabetes, diagnosed hypertension, cancer, asthma, chronic obstructive pulmonary disease, and renal disease), biomarkers (total cholesterol, high-density lipoprotein cholesterol, low-density lipoprotein cholesterol, c-reactive protein, creatinine), number of hospital admissions in last year, and prescribed non-oral
glucocorticoids; and time-variant use of disease-modifying anti-rheumatic drugs and non-steroidal anti-inflammatory drugs; the practice identifier was included as a random intercept to account for clustering effect. \*These estimates were additionally adjusted for the type of immune-mediated inflammatory disease diagnosed

We defined several time-variant glucocorticoid variables to quantify current and cumulative drug exposure: i) ever use from one year prior to follow-up start (binary variable); ii) current daily use (i.e. whether or not the patient was prescribed glucocorticoids at a given time point [binary variable]); iii) current daily dose per 5 mg/day with zero value when medication was not prescribed (continuous and categorical variables: non-use, 1-4.9 mg, 5.0-14.9 mg, 15.0-24.9 mg, ≥ 25.0 mg/day); iv) cumulative dose since one year prior to follow-up start per 1,000 mg (i.e. sum of the total dose prescribed up to that point divided by 1,000; considered as continuous and categorical variables: non-use, 1-959 mg, 960-3,054 mg, 3,055-7,299 mg, and ≥7,300 mg; as defined previously[19, 20]).

### Outcome measures

The primary outcome was the first occurrence of a composite of fatal and non-fatal cardiovascular diseases (all-cause CVD). Secondary outcomes were the first occurrence of the following common types of CVDs: atrial fibrillation, heart failure, myocardial infarction, cerebrovascular disease, peripheral arterial disease and abdominal aortic aneurysm. Diagnostic codes[21] used to define the outcomes are listed in Table 3 in S1 File and have been validated and used in multiple previous studies[22-26].

### Confounding variables

We considered the following variables as a priory confounders: baseline age, sex, ethnicity, socioeconomic status (index of multiple deprivation[24, 27], area-based indicator linked through the patient’s home postcode), smoking status, body mass index (BMI), biomarkers (total, high and low density lipoprotein-cholesterol, systolic blood pressure, c-protein reactive protein, creatinine), underlying disease (e.g. rheumatoid arthritis), comorbidities recorded in primary or hospital care (diabetes, diagnosed hypertension, cancer, asthma, chronic obstructive pulmonary disease [COPD], renal disease); prescribed non-oral glucocorticoid medication (inhaled, nasal, parenteral/intra-articular, topical and rectal), and the number of hospital visits 1 year before baseline. We also considered time-variant prescribed medication (disease-modifying antirheumatic drugs and non-steroidal anti-inflammatory drugs) during follow-up. Detailed definition of covariates are shown in Text 1 in S1 File

### Statistical analysis

We replaced missing daily dose of oral glucocorticoids (i.e. during tapering periods) and confounders through multiple imputation with chained equations with generation of 25 datasets (Text 1 in S1 File). Models for dose imputation included patient demographics (i.e. age, sex, and index of multiple deprivation[28]), underlying immune-mediated inflammatory disease, time between follow-up start and prescription, type of oral glucocorticoid (e.g. prednisone) and diagnosed comorbidities (e.g. diabetes).

We used standard descriptive statistics to describe baseline patient characteristics. We estimated cumulative probabilities of CVD outcomes using Kaplan-Meier methods. We calculated incident rates with 95% confidence intervals (95% CI) dividing the number of patients with incident CVD by the total number of person-years of follow-up.

We assessed the association between the outcomes and each of the oral glucocorticoid exposures using Cox proportional hazards models adjusted for the a priori confounders, with the practice identifier included as a random intercept to account for clustering effect. No interaction terms were included. The proportional hazards assumption was assessed using Schoenfeld residuals tests. The primary analysis was based on covariate imputed data. We generated models for each of the 25 imputed datasets and pooled estimates and accompanying 95% CIs following Rubin’s rules. We used 2-sided tests and considered significant at p<0.05. We performed the data management in Stata (StataCorp LP, College Station, USA; version 15) and analyses in R (http://cran.r-project.org/; version 3.3.1).

In secondary analyses, we modelled cardiovascular risk separately for men and women, for each of the 6 immune-mediated inflammatory diseases studied, and according to duration of these diseases at the start of follow-up (newly diagnosed/incident, within 2 years and over 2 years since diagnosis).

In sensitivity analyses, we obtained estimates from complete case models (i.e. restricted to patients with complete covariate data), from models including covariates with a separate category for missing data and from models unadjusted for biomarker data with a level of missingness >60%. We also additionally adjusted the models for the level of disease activity. We defined periods of active disease based on c-reactive protein and erythrocyte sedimentation rate levels (≥10 mg/mL and ≥30 mm/h, respectively) and the glucocorticoid daily dose (increase in prednisolone-equivalent dose by >5 or 10 mg that was sustained for over 3 weeks) (Text 1 in S1 File).

## RESULTS

### Patient characteristics

The study included 87,794 adults from 389 general practices with at least one immune-mediated inflammatory disease diagnosed; 25,581 with polymyalgia and/or giant cell arteritis, 27,739 (31.6%) with inflammatory bowel disease, 25,324 (28.8%) with rheumatoid arthritis, 5,199 (5.9%) with vasculitis, and 3,951 (4.5%) with systemic lupus erythematosus (Table 1). The overall mean age was 56 years (SD 18.3), 29,935 (34.1%) were men and 21,264 (24.2%) were current smokers. At baseline, the mean duration since immune-mediated disease diagnosis was 9.6 years (SD=8.7; range from 6.8 years for polymyalgia and/or giant cell arteritis and 11.7 years for inflammatory bowel disease). The most common patient comorbidities were hypertension (25.1%), asthma (14.6%) and diabetes (6.4%).

In the year prior to follow-up start, 14,356 (16.4%) patients were prescribed oral glucocorticoids, 13,331 (15.2%) inhaled or nasal glucocorticoids and 39,690 (45.2%) non-steroidal anti-inflammatory drugs. During follow-up 18,877 (21.5%; range 4.2% for polymyalgia and/or giant cell arteritis to 48.0% for rheumatoid arthritis) received disease-modifying antirheumatic drugs.

### Incidence of fatal and non-fatal cardiovascular diseases

The median time of follow-up per patient was 5.0 (IQR 2.0-6.2) years. A total of 13,426 incident cardiovascular events occurred (15.3% of patients) over 541,655 person-years of follow-up (Table 4 in S1 File), including 6,013 episodes of atrial fibrillation, 4,727 of heart failure and 2,809 of acute myocardial infarction. The incidence of all-cause CVD was 24.8 per 1,000 person-years (95%CI 24.4-25.2). It increased from 18.5 (95%CI 18.1-18.9) per 1,000 person-years for periods of non-glucocorticoid use to 45.6 (95%CI 42.1-49.2) for periods of ≥25mg daily dose; and from 19.9 (95%CI 19.3-20.5) for unexposed periods to 26.4 (95%CI 25.5-27.2) per 1,000 person-years for ≥7,300 mg cumulative those). A total of 7,940 cardiovascular events happened during periods of non-exposure.

### Cumulative probabilities of cardiovascular diseases

The Kaplan-Meier estimates of all-cause CVD at 1 year increased from 1.5% (95%CI 1.4%-1.6%) for periods of non-use, through 3.8% (95%CI 3.4%-4.3%) for <5 mg, to 9.1% (95%CI 7.6%-10.7%) for ≥25.0 mg daily dose; and from 7.6% (95%CI 7.4%-7.8%) for unexposed periods to 10.3% (95%CI 9.7%-10.9%) for ≥7,300 mg cumulative dose (Table 5 in S1 File). We found higher dose-response estimates in men than in women, in women with systemic lupus erythematosus, men with rheumatoid arthritis, and for atrial fibrillation and heart failure (Tables 6&7 in S1 File).

### Relationship between glucocorticoid dose and cardiovascular diseases

The increase in the hazard of all-cause CVD per 5 mg increase in daily dose was 1.08 (95%CI 1.07-1.10 per 5 mg/day), ranging from 1.07 (95%CI 1.06-1.09) for inflammatory bowel disease to 1.30 (95%CI 1.22-1.38) for systemic lupus erythematosus (Table 2). We found strong dose-response estimates for current daily doses of <5.0mg for all immune-mediated diseases (HR=1.74, 95%CI 1.64-1.84; range 1.52 for polymyalgia and/or giant cell arteritis to 2.82 for systemic lupus erythematosus), for all cardiovascular outcomes and for daily and cumulative dose (Figures 1-3; Figures 2-13 in S1 File). The highest glucocorticoid dose-response estimates were for heart failure and for acute myocardial infarction. We found similar patterns in sensitivity analyses, including restriction to patients with complete covariate data (Tables 8&10 in S1 File). Daily and cumulative dose-response estimates were generally higher amongst patients with longer underlying inflammatory disease duration and in those newly diagnosed (Tables 11&14 in S1 File). Further, adjustment for the level of disease activity generally decreased the dose-response estimates, but associations remained statistically significant (Tables 15&16 in S1 File).

**Fig 2.**
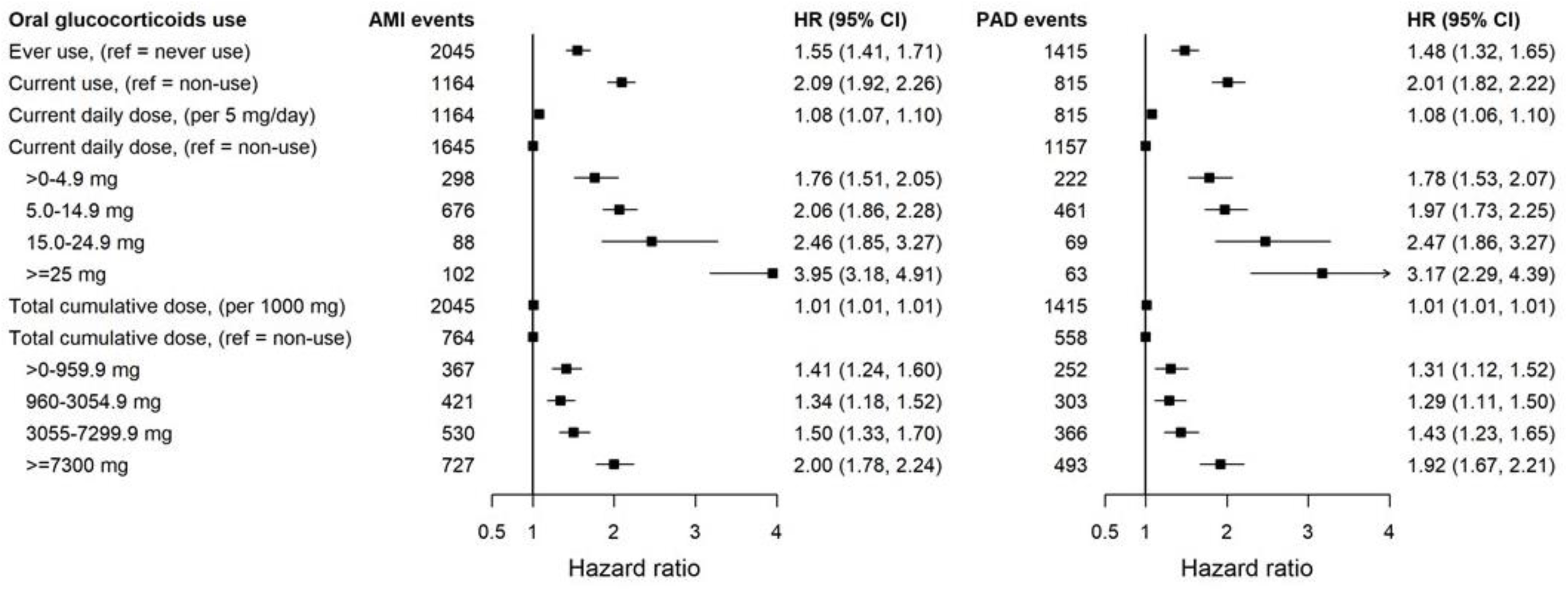
Associations between time variant oral glucocorticoid prednisolone-equivalent dose and incident acute myocardial infarction and peripheral arterial disease for patients with 6 immune-mediated inflammatory diseases. Note: AMI, acute myocardial infarction; CI, confidence interval; HR, Hazard ratios from Cox proportional imputed models adjusted for baseline age, sex, index of multiple deprivation, smoking status, ethnicity, body mass index, type of immune-mediated inflammatory disease, comorbidities (diabetes, diagnosed hypertension, cancer, asthma, chronic obstructive pulmonary disease, and renal disease), biomarkers (total cholesterol, high-density lipoprotein cholesterol, low-density lipoprotein cholesterol, c-reactive protein, creatinine), number of hospital admissions in last year, and prescribed non-oral glucocorticoids; and time-variant use of disease-modifying anti-rheumatic drugs and non-steroidal anti-inflammatory drugs; the practice identifier was included as a random intercept to account for clustering effect. PAD, peripheral arterial disease.

**Fig 3.**
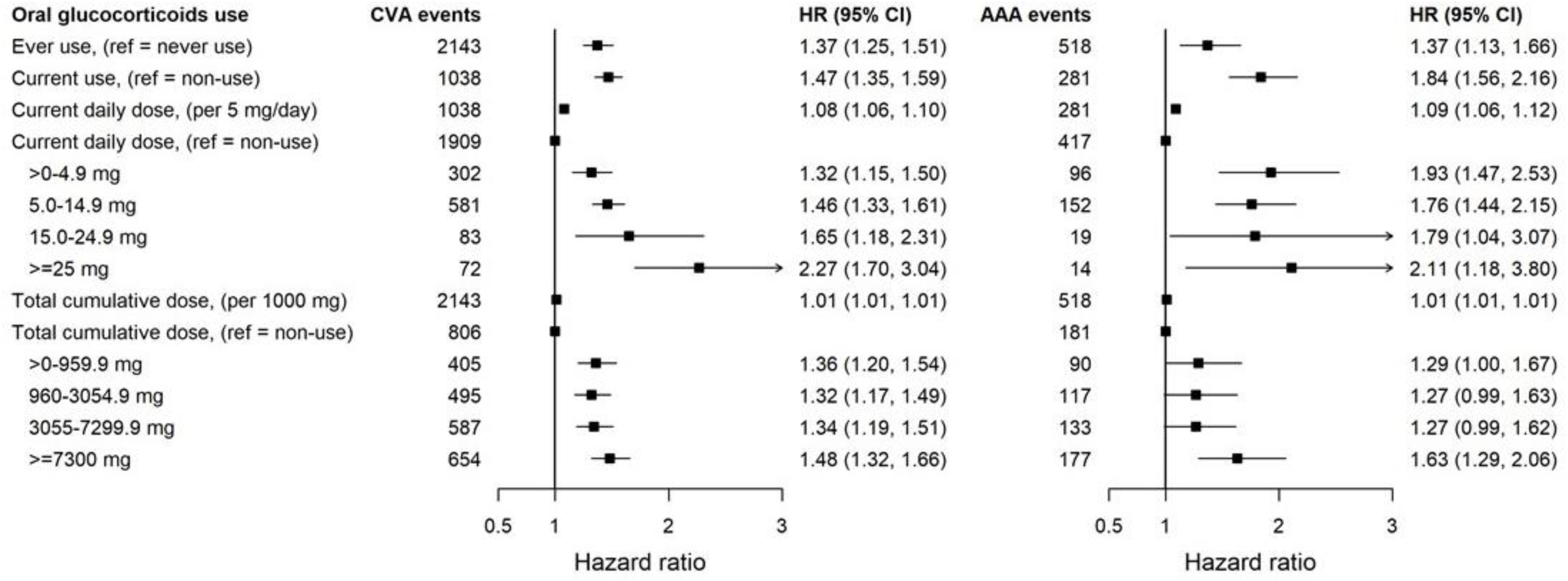
Associations between time variant oral glucocorticoid prednisolone-equivalent dose and incident cerebrovascular disease and abdominal aortic aneurysm for patients with 6 immune-mediated inflammatory diseases. Note: AAA, abdominal aortic aneurysm; CI, confidence interval; CVA, cerebrovascular disease; HR, Hazard ratios from Cox proportional imputed models adjusted for baseline age, sex, index of multiple deprivation, smoking status, ethnicity, body mass index, type of immune-mediated inflammatory disease, comorbidities (diabetes, diagnosed hypertension, cancer, asthma, chronic obstructive pulmonary disease, and renal disease), biomarkers (total cholesterol, high-density lipoprotein cholesterol, low-density lipoprotein cholesterol, c-reactive protein, creatinine), number of hospital admissions in last year, and prescribed non-oral glucocorticoids; and time-variant use of disease-modifying anti-rheumatic drugs and non-steroidal anti-inflammatory drugs; the practice identifier was included as a random intercept to account for clustering effect.

## DISCUSSION

In this longitudinal study of 87,794 adults diagnosed with at least one of six common immune-mediated inflammatory diseases, we quantified oral glucocorticoid dose-dependent risks of all-cause and type-specific CVDs taking into account changes in prescribed medication over time. At 1 year, the cumulative risk of all-cause CVD increased from 1.5% during periods without medication, through 3.8% for a daily prednisolone-equivalent dose <5mg, to 9.1% for periods with a daily dose of ≥25.0mg. We found strong dose-dependent increases in hazards of all-cause CVD, atherosclerotic diseases, heart failure, atrial fibrillation and abdominal aortic aneurysm, regardless of the underlying immune-mediated disease, its activity and duration. The cardiovascular risk profile of the study patients showed high prevalence of modifiable risk factors, including current smoking (24.2% of patients), BMI ≥30 kg/m^2^ (24.5%) and hypertension (25.1%).

Previous studies have reported increased risk of composites of cardiovascular[2, 3, 11, 12] or coronary heart disease[1], myocardial infarction[2, 3, 11, 14, 15], heart failure[2, 3, 11], stroke[2, 3, 16] and atrial fibrillation[7, 13] in current glucocorticoid users. Some found an increased risk of CVD only for daily doses of 5-10mg or higher[13, 16, 17]. Estimates from previous studies were based on current, baseline medication use[2, 3, 7, 11-16] or dose, or average glucocorticoid dose in the last 6-12 months[1, 3], without consideration of previously administered doses and changes in dose or medication use over time. Consistent with our findings, other studies assessing the relationship between glucocorticoid use and the risk of different types of CVDs reported stronger associations for heart failure than for other cardiovascular outcomes[2, 3].

The high absolute risk of CVD in patients receiving high doses of glucocorticoids, of similar magnitude to that of patients with diabetes or established CVD, warrants the need to implement and evaluate intensive lifestyle modification interventions to this high-risk group. The dose-dependent increased risk of CVDs, including atherosclerotic diseases, heart failure and atrial fibrillation, observed in our study, supports the need for close monitoring of cardiovascular risk in patients diagnosed with immune-mediated inflammatory diseases during glucocorticoid treatment and in the period after therapy discontinuation. Of cardiovascular risk scores currently used to guide decision on when to start primary cardiovascular prophylaxis, only the QRISK3[29] considers whether the patient is currently taking glucocorticoids (as a binary ‘yes/no’ predictor that ignores dose and recent exposure) and whether he/she is diagnosed with rheumatoid arthritis or systemic lupus erythematosus. Further refinements of this risk prediction tool, taking into account cumulative and/or current dose, might therefore improve its performance to identify patients in need for primary cardiovascular prevention. Our findings also emphasise the importance of rapid glucocorticoid dose tapering and discontinuation as soon as disease control is achieved, as well as the importance of evaluating the safety profile of alternative therapeutic options for patients with autoimmune-mediated inflammatory diseases.

The estimation of drug dose-response risks in this population-based cohort of all people with immune-mediated inflammatory diseases with different levels of activity and duration minimised the introduction of selection bias and increased the generalisability of the results. The use of linked health data from primary care and hospital facilities and the mortality registry, and diagnostic codes extensively used and validated for cardiovascular research[23-26, 30, 31], increased ascertainment of all the outcomes assessed. Information on prescribed medication is prospectively collected and includes all prescriptions issued in primary care, where patients with immune-mediated diseases are primarily treated. We derived the dose of oral glucocorticoids and the duration of prescribed medication from the directions given to patients on how to take their treatment. During periods of dose tapering, when these directions were unspecific (e.g. written ‘as directed’), we used the longitudinal doses prescribed to the patients to impute the dose taken. The lack of data on hospital prescribed medication and on drug adherence is likely to have resulted in underestimation of the dose taken when specialists treated the patients and might have overestimated the dose taken in periods of low disease activity for some patients. The resulting misclassification is likely to have reduced the size of dose-response estimates. We minimised time-related bias through use of time-variant medication variables (both exposure and confounders) and a start of patient follow-up that was unrelated to the start or use of glucocorticoid therapy. Although the main purpose of the study was to provide estimates of oral glucocorticoid dose-response for patients with the inflammatory diseases studied without making aetiological inferences, we examined the effect of confounding by indication and ascertainment bias through adjustment by periods of disease activity and disease-modifying antirheumatic drugs use, and performing analyses according to duration of the underlying disease. Dose-response associations remained strong and statistically significant. We adjusted estimates of risk for established cardiovascular risk factors (e.g. smoking, hypertension and diabetes), concomitant use of medications (e.g. time-variant disease-modifying antirheumatic drugs, non-steroidal anti-inflammatory drugs, non-oral glucocorticoid use). In the primary analysis, we used multiple imputation to handle missing baseline biomarkers and smoking data. We found similar patterns of dose-response in sensitivity, including analyses in which we used a separate category for missing covariate data, in those restricted to individuals with complete covariate data and in those unadjusted for baseline biomarker information with high level of missingness (>60% of patients).

In conclusion, we reported improved estimates of dose-dependent risks of CVDs. Our findings highlight the importance of implementing and evaluating targeted intensive cardiovascular risk factor modification interventions; promptly and regularly monitor patient cardiovascular risk, beyond diagnosis of inflammatory arthropathies and systemic lupus erythematosus, even when prescribing low prednisolone-equivalent doses; and the need for refining existing risk prediction tools for primary prevention of CVDs. Furthermore, the estimates of risk can be used to conduct cost-effectiveness and benefit-harm evaluations that guide the introduction of newly licensed glucocorticoid-sparing drugs for the treatment of immune-mediated inflammatory diseases. These estimates are to be complemented by future work on the estimation of risk of cardiovascular events beyond the first occurrence of CVD that are considered in calculations of glucocorticoid-associated costs.

## Data Availability

Access to raw data can be requested from the CPRD (https://cprd.com)

## ACKNOWLEDGEMENTS

AWM is supported by the Medical Research Council TARGET Partnership Grant (Treatment According to Response in Giant cEll arTeritis) (MR/N011775/1), the National Institute for Health Research (NIHR) infrastructure @ Leeds, including the NIHR Biomedical Research Centre (IS-BRC-1215-20015), and NIHR Medtech and In vitro Diagnostics Co-operatives (MIC-2016-015). JW is supported by the NIHR infrastructure @ Leeds. The views expressed are those of the author(s) and not necessarily those of the NHS, the NIHR or the Department of Health and Social Care. The study funders had no role in the study design, data collection, analysis or interpretation, in the writing of the paper or in the decision to submit the paper for publication.

## SUPPORTING INFORMATION

**S1 File. Supplemental material**

